# Early COVID-19 vaccine effectiveness of XBB.1.5 vaccine against hospitalization and ICU admission, the Netherlands, 9 October - 5 December 2023

**DOI:** 10.1101/2023.12.12.23299855

**Authors:** C. Henri van Werkhoven, Anne-Wil Valk, Bente Smagge, Hester E. de Melker, Mirjam J. Knol, Susan J.M. Hahné, Susan van den Hof, Brechje de Gier

## Abstract

We present early vaccine effectiveness (VE) estimates of the 2023 seasonal COVID-19 vaccination campaign using XBB.1.5 vaccine against COVID-19 hospitalization and ICU admission in previously vaccinated adults ≥60 years old in the Netherlands. We compared vaccination status of 2050 hospitalizations including 92 ICU admissions with age group-, sex-, region- and date-specific population vaccination coverage between 9 October and 5 December 2023. VE against hospitalization was 70.7% (95% CI: 66.6; 74.3), VE against ICU admission was 73.3% (95% CI: 42.2; 87.6).

---

In the Netherlands, the 2023 seasonal COVID-19 vaccination campaign started on 2 October, targeting persons aged 60 years and older, healthcare workers, pregnant women and medical risk groups using monovalent XBB1.5 Comirnaty vaccine. All residents of the Netherlands aged 60 years and older received a personal invitation for vaccination by mail between 2 October and the end of November. Up to 3 December 2023, 42.9% of residents aged 60 and older had received the 2023 seasonal COVID-19 vaccination [1]. The seasonal vaccination campaign runs until 22 December 2023. To inform the public and policy makers, we here provide an early estimate of the vaccine effectiveness (VE) of the 2023 seasonal vaccination campaign against COVID-19 hospitalization and ICU admission.

## Data sources and method

We used the screening method to estimate the 2023 seasonal VE among persons aged 60 years and older (birth year 1962 or before) with at least one previous COVID-19 vaccination, included in the population register of the Netherlands on 25 September 2023. Hospitalizations with admission dates between 9 October and 5 December 2023 were extracted from the NICE COVID-19 database on 11 December 2023, to account for registration delay. This dataset contained around 55% of all COVID-19 hospitalizations during the study period in the Netherlands, compared to anonymous data from the National Coordination Center for Patient Distribution [2]. Hospitalization data were linked deterministically based on Citizen Service Number to the national COVID-19 vaccination database (CIMS). COVID-19 vaccinations are registered in CIMS when vaccinees provide informed consent for registration. The percentages of persons vaccinated at the Municipal Health Services consenting to registration in CIMS was ≥95% during previous booster campaigns, and 98% for the 2023 seasonal campaign [1]. We included only persons with at least one previous vaccination registered in CIMS since January 2021, based on the assumption that this population is most likely to consent to registration of 2023 seasonal vaccination as well, thereby minimizing misclassification [3]. While formally the campaign started on 2 October 2023, some facilities started vaccination on September 25. Therefore, we included vaccinations from 25 September onward as 2023 seasonal doses. Persons who received a COVID-19 vaccination in the 90 days before 25 September were excluded from the analysis. To take into account the delay in immunological response after vaccination, a person’s vaccination status changed to “seasonal dose received” 7 days after administration of the dose. The 7 days between vaccination and the status “seasonal dose received” were excluded from the analysis. Vaccination status of the cases on the date of admission was determined in the same way. The CIMS registry was used to calculate the proportion of the population with seasonal dose received, stratified by the following covariates: calendar date, sex, geographic region (25 levels) and 5-year age group. Persons contributed to the proportion of the population vaccinated until the 15^th^ of the month of death or emigration or end of the study period.

VE and 95% confidence intervals (95%CI) were estimated using a logistic regression model, with vaccination status as dependent variable and the covariate-specific logit of seasonal dose received in the population as offset. The exponentiated intercept of this model is interpreted as the relative risk (RR), and VE is calculated as (1-RR) x 100% [4].

## VE estimates

After exclusion of 86 hospitalizations (including 3 ICU admissions) occurring in the 7 days after administration of the seasonal dose, we included 2,050 hospitalized persons, of whom 295 (14.4%) had received the 2023 seasonal COVID-19 vaccination (Table 1). The number of hospitalizations was higher among persons aged 75 years and older compared to persons aged 60-74 years (Figure 1). Towards the end of the study period, more seasonal vaccinees were hospitalized, reflecting the gradual roll-out of the campaign (Figure 2). VE against hospitalization was estimated at 70.7% (95%CI: 66.6;74.3, Table 1). Of the included hospitalizations, 92 concerned ICU admissions, with an estimated VE of 73.3% (95%CI 42.2;87.6). The VE against hospitalization was slightly lower for the age group 60-74 years (68.3%, 95%CI 58.3;75.9) than for persons aged 75 years and older (71.4%, 95%CI 66.7;75.4). A year ago, for the period 3 October 2022-12 December 2022 we estimated a slightly lower VE of 64% (95%CI 59-68%) of bivalent booster vaccination against hospitalization for the age group 60-79 years, although the 2022 autumn campaign was closer in time to the previous campaign in spring 2022 [5].

**Table 1.** COVID-19 hospitalizations and ICU admissions included in the analysis by 2023 seasonal vaccination status and estimated vaccine effectiveness (VE) adjusted for calendar date, region (25 levels), sex and 5-year age group, the Netherlands, 9 October-5 December 2023.

| Outcome | Age group | N cases with 2023 seasonal vaccination | N cases without 2023 seasonal vaccination | VE (95% CI) |
| --- | --- | --- | --- | --- |
| **COVID-19 hospitalization** | 60+ | 295 | 1755 | 70.7%<br>(66.6;74.3) |
|  | 60-74 | 59 | 681 | 68.3%<br>(58.3;75.9) |
|  | 75+ | 236 | 1074 | 71.4%<br>(66.7;75.4) |
| **COVID-19 ICU admission** | 60+ | 8 | 84 | 73.3%<br>(42.2;87.6) |

**Figure 1.**
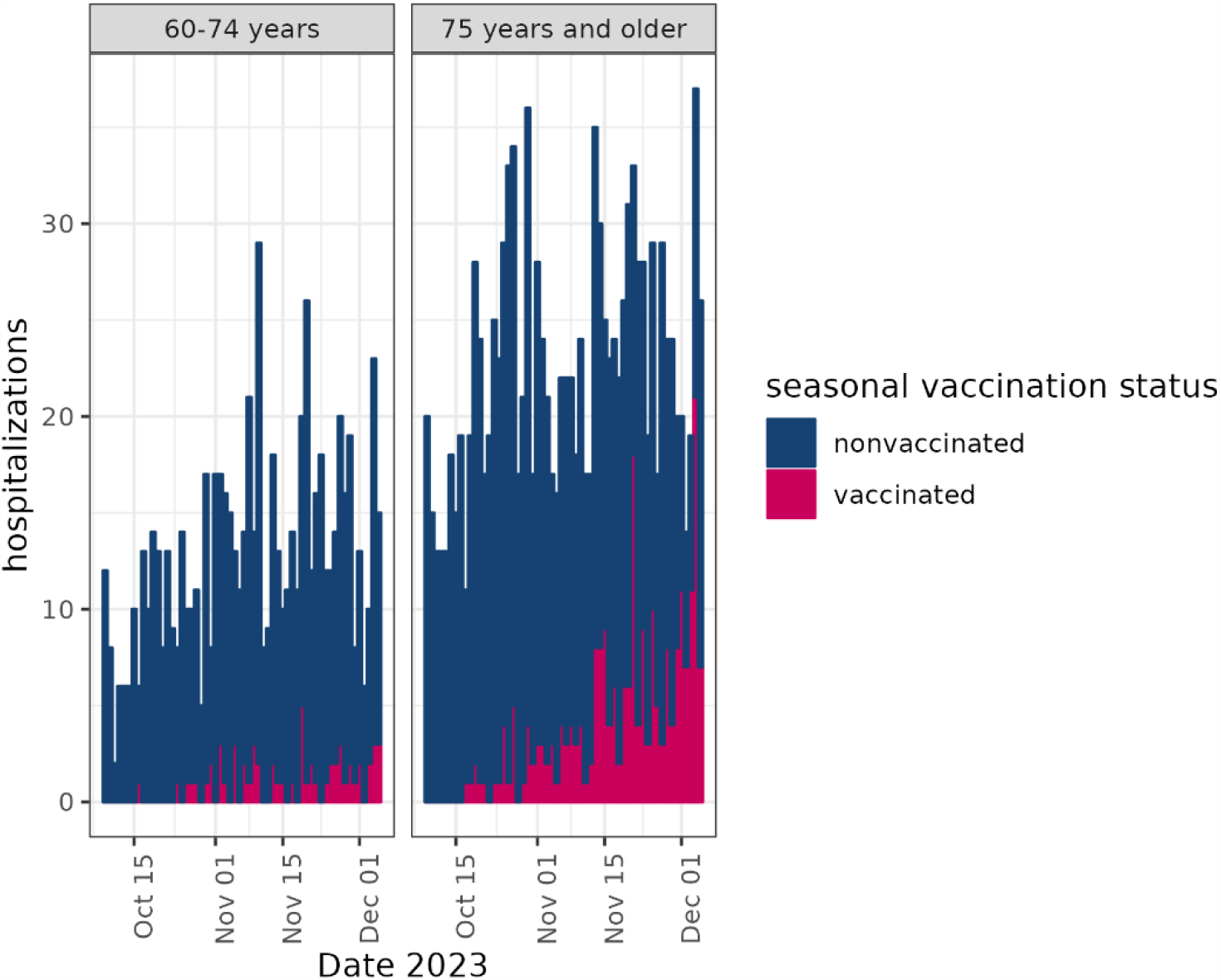
COVID-19 hospitalizations and ICU admissions included in the analysis per date, seasonal vaccination status and age group, the Netherlands, 9 October – 5 December 2023.

**Figure 2.**
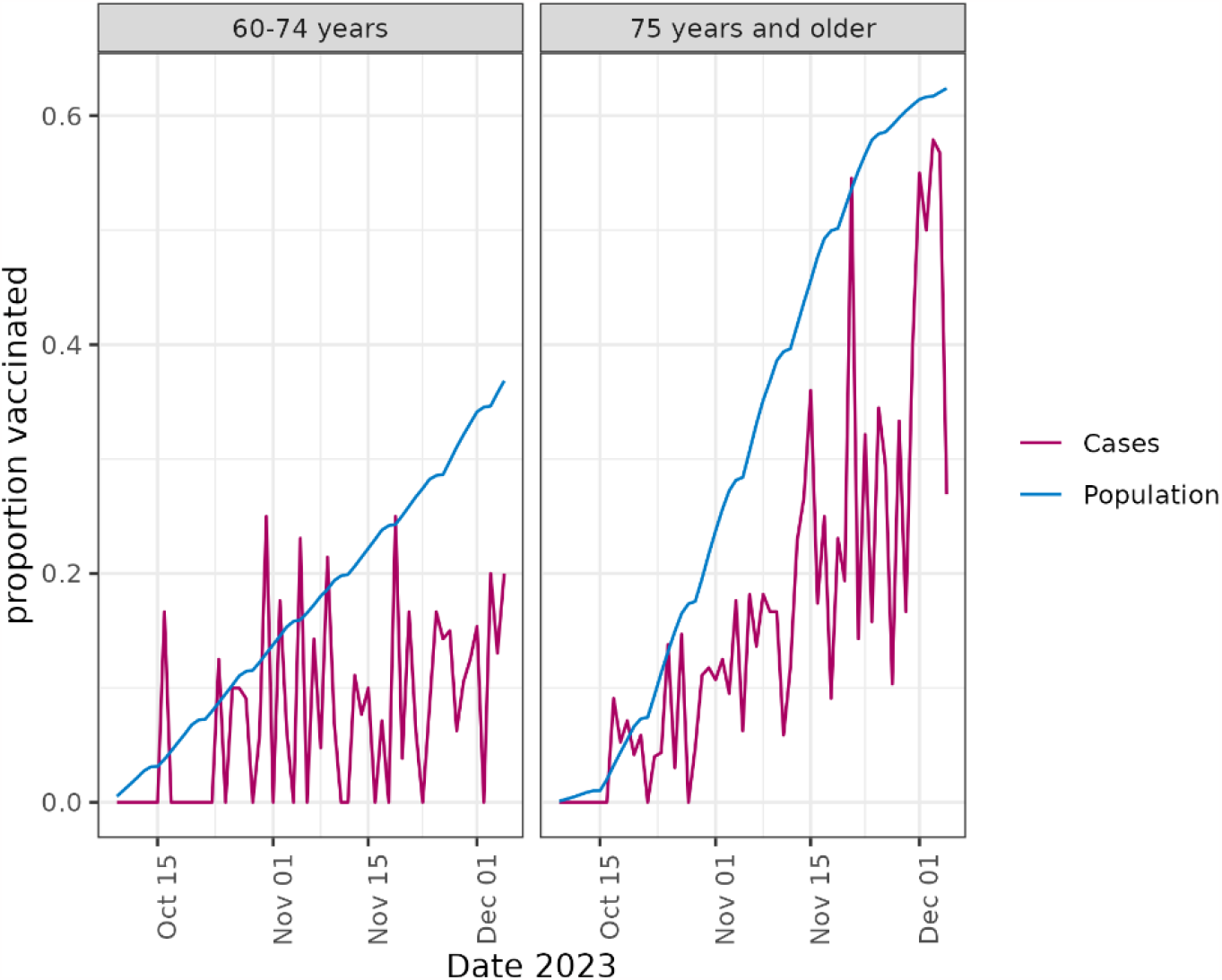
Proportion of seasonal vaccinees among hospitalized cases and among the population aged 60 years and older eligible for a booster vaccination on 25 September 2023, by date and age group, the Netherlands, 9 October – 5 December 2023.

We used the screening method, which we deemed more appropriate than a cohort approach because only a proportion of cases is known. While the NICE hospitalization database was complete in previous years, this is no longer the case as due to registration burden not all hospitals still contribute data. To adjust for possible geographic differences in completeness of hospitalization data, we included region in our model.

A number of limitations may have led to an underestimation of the VE. We were not able to adjust the estimates for comorbidities; such an adjustment generally leads to a higher VE estimate [6]. A study from Denmark reported an early VE estimate against COVID-19 hospitalization adjusting for comorbidities of 75.3% between 8 and 26 October, among persons aged 65 and older [7]. As our VE estimate is based on the first period of the seasonal campaign (see Figure 2), especially the beginning of the study period will represent the VE among early vaccinees, where frail persons could be overrepresented. Further, we defined the vaccination status of cases on the date of hospitalization, which will be some days after infection and disease progression. Because vaccination coverage was quickly changing during our study period, a relevant proportion of cases was recently vaccinated and might have lacked time for the vaccine to prevent severe disease. If we assess vaccination status of cases 7 days before hospital admission, VE is estimated at 74% (95%CI 69.6;77.7) against hospitalization for ages 60 and older.

Conversely, our VE might be biased upward due to a better general health of seasonal vaccinees compared to the reference group (healthy vaccinee bias). In the Danish study, an HR of 0.851 was found for hospitalizations by other causes than COVID-19 [7]. In a previous study by our group on VE against COVID-19 mortality, we found a high VE against all-cause mortality, indicating a likely healthy vaccinee bias [6].

For the highest risk groups in the Netherlands, the 2023 seasonal campaign constitutes the fifth booster dose offered after primary vaccination. However, for only 0.2% of the population aged 60 and older having received the 2023 seasonal dose this pertained a fifth booster. For the majority (76%), the 2023 seasonal dose was the fourth booster dose. On 24 September 2023, the median time since the previous booster dose was 50 weeks, with an interquartile range (IQR) of 48 to 52 weeks. The number of previous booster doses was lower among the study population not (yet) having received the 2023 seasonal vaccine (33% 1, 27% 2 and 24% 3 previous boosters), and on 24 September 2023 the median time since the last booster dose was 61 weeks (IQR 49-90 weeks). If a higher number of doses and/or a shorter time since the last vaccination still conferred some protection during our study period, this may have led to an overestimation of the VE. However, a study among 6 European countries found that the number of booster doses did not have a relevant effect on VE against hospitalization, and VE had generally waned 6 months after any booster dose [8].

## Conclusion

This early estimate suggests a high VE against hospitalization and ICU admission in the first two months of seasonal COVID-19 vaccination with XBB.1.5 vaccine. The VE is expected to decrease as the time since vaccination increases in the coming months, as observed in previous COVID-19 vaccination campaigns in Europe [9, 10]. Still, it is likely that seasonal COVID-19 vaccination is an effective intervention for reducing the burden of severe COVID-19 during the coming winter months, next to the reduction already achieved in the first few months. An increase in uptake of the seasonal vaccine could further enhance its public health benefit.

## Data Availability

Data are not publicly available due to privacy law.

## Ethical statement

The Centre for Clinical Expertise at the RIVM assessed the research proposal (reference EPI-520). They verified whether the work complies with the specific conditions as stated in the law for medical research involving human subjects (WMO), and were of the opinion that the research does not fulfill one or both of these conditions and therefore conclude it is exempted for further approval by the ethical research committee.

## Funding statement

This work was funded by the Dutch Ministry of Health, Welfare and Sports.

## Acknowledgements

The authors would like to acknowledge Nicolette de Keizer and the NICE R&S team.

## Conflicts of Interest

CHvW reports financial and non-financial research support from DaVolterrra and bioMérieux; financial research support from LimmaTech; consultancy fees from MSD and Sanofi-Pasteur (all payments to the University Medical Centre Utrecht). Other authors have nothing to report.

## Authors’ contributions

Conceptualization: BdG, CHvW. Data analysis: CHvW, BdG. First draft of manuscript: BdG. Critical discussion and revision of manuscript: A-WV, BS, MJK, SJMH, HEdM, SvdH. All authors have read and approved the final manuscript.

## Notes

### Competing Interest Statement

CHvW reports financial and non-financial research support from DaVolterrra and bioMerieux; financial research support from LimmaTech; consultancy fees from MSD and Sanofi-Pasteur (all payments to the University Medical Centre Utrecht). Other authors have nothing to report.

